# Correlation between times to SARS-CoV-2 symptom onset and secondary transmission undermines epidemic control efforts

**DOI:** 10.1101/2021.08.29.21262512

**Authors:** Natalie M. Linton, Andrei R. Akhmetzhanov, Hiroshi Nishiura

**Affiliations:** Kyoto University School of Public Health, Yoshidakonoe-cho, Sakyo-ku, Kyoto city, 606-8501, Japan; Graduate School of Medicine, Hokkaido University, Kita 15 Jo Nishi 7 Chome, Kita-ku, Sapporo-shi, Hokkaido 060-8638, Japan; College of Public Health, National Taiwan University, 17 Xu-Zhou Road, Taipei 10055, Taiwan; Core Research for Evolutional Science and Technology (CREST), Japan Science and Technology Agency, Saitama, Japan

## Abstract

Severe acute respiratory coronavirus 2 (SARS-CoV-2) infections have been associated with substantial presymptomatic transmission, which occurs when the generation interval—the time between infection of an individual with a pathogen and transmission of the pathogen to another individual—is shorter than the incubation period—the time between infection and symptom onset. We collected a dataset of 257 SARS-CoV-2 transmission pairs in Japan and jointly estimated the mean generation interval (3.7–5.1 days) and mean incubation period (4.4–5.7 days) as well as measured their dependence (Kendall’s tau of 0.4–0.6), taking into consideration demographic and epidemiological characteristics of the pairs. The positive correlation between the two parameters demonstrates that reliance on isolation of symptomatic COVID-19 cases as a focal point of control efforts is insufficient to address the challenges posed by SARS-CoV-2 transmission dynamics. Accounting for this dependence within SARS-CoV-2 epidemic models can also improve model estimates.

## Introduction

The generation interval and incubation period of an infectious disease are key epidemiological parameters used to inform outbreak response. The former describes the infectiousness of a pathogen in a host—it represents the time between when a host (an infector) is infected with a pathogen and when they transmit that pathogen to another host (an infectee). In contrast, the incubation period represents the time between infection and development of symptomatic disease. It reflects the pathogen replication rate and provides a basis for predicting prognosis. It also indicates how long an infection may remain unnoticed in an individual, and thus for emerging diseases it is used to determine quarantine periods.^1,2^ Together, the generation interval and incubation period provide insight into the intrinsic dynamics of infection and characterize the effectiveness of public health interventions on its control.

At the beginning of the coronavirus 2019 (COVID-19) pandemic, the mean incubation period of severe acute respiratory disease coronavirus 2 (SARS-CoV-2)—the pathogen causing COVID-19—was rapidly estimated.^1–4^ Estimates of the mean serial interval, which is the time between symptom onset in an infector and symptom onset in a person they infect, quickly followed.^5–8^ However, due to difficulty in ascertaining exposure times of cases and directionality of transmission between epidemiologically linked cases, few attempts were made to estimate the generation interval.^6–10^ Instead, the serial interval was often used as a proxy for the generation interval when estimating epidemiological quantities. For example, it has been used to estimate the basic reproduction number—the average number of persons infected by a single infector in a completely susceptible population—and the effective reproduction number—the average number of persons infected by a single infector in the presence of existing infections and interventions.^11,12^ However, use of the serial interval as a proxy for generation time can lead to biased estimates of the effective reproduction number due to factors such as differences in their variances, the presence of asymptomatic infections, and the ability for the serial interval to have negative values (infectee onset preceding infector onset).^8,13–16^

Generation intervals depend on many factors, such as the ratio of infected to susceptible persons among contacts, as well as the behavior of infected persons.^17^ Actions such as self-isolation after symptom onset can shorten generation intervals by limiting the opportunity for infected individuals to infect others during their early symptomatic period. However, if an infected person can transmit the pathogen before symptom onset, then isolation of symptomatic persons alone is insufficient to control the spread.^18^ For pathogens such as SARS-CoV-2, interventions targeting nonsymptomatic cases appears vital^19^ due to the large fraction of presymptomatic and asymptomatic transmission.^7,20,21^

Frequently, estimates of the generation interval of SARS-CoV-2 have been derived from the serial interval and formulated following implicit (and unsupported) assumptions that: i) there is no asymptomatic transmission,^22^ ii) the incubation period and generation interval are independent.^23^ However, these assumptions are clearly flawed with respect to SARS-CoV-2 transmission. In the case of asymptomatic transmission, there is substantial evidence of transmission from asymptomatic infectors,^20,24^ as well as plenty of asymptomatic cases demonstrating the existence of asymptomatic and presymptomatic infectees. Likewise, correlation between the generation interval and incubation period of SARS-CoV-2 was shown to be biologically plausible, as evidenced by the peak viral load of SARS-CoV-2 occurring around the time of symptom onset.^25^ However, previous studies that considered such a correlation did not attempt to directly estimate it.^26^

In this study, we provide direct evidence of their correlation and jointly estimate the generation interval and incubation period using transmission pairs identified in Japan in 2020. We assessed whether either interval or their correlation varied based on demographic and epidemiological characteristics. Accounting for correlation between the generation interval and incubation period can considerably help quantify SARS-CoV-2 transmission and improve characterization of the effectiveness of public health interventions by preventing underestimation of the proportion of presymptomatic transmission and the effect of isolation of symptomatic cases on epidemic control.

## Methods

### Ethics oversight

This study was approved by the Medical Ethics Board of the Graduate School of Medicine at Kyoto University (R2676). It uses data published online by public health jurisdictions in Japan.

### Study population and setting

We compiled a dataset of COVID-19 transmission pairs using openly published case data from reporting jurisdictions (prefectures and cities) in Japan, focusing on detecting pairs for whom directionality of transmission could be determined with some degree of certainty. Cases were limited to those reported during the calendar year 2020. Jurisdiction reporting practices changed over time, with details generally becoming sparser over time, as concerns grew around infection-related stigmatization,^40^ as well as in prefectures with large case loads.

Among the information publicly shared were links between cases and links to common exposures (e.g., a medical facility, event, or restaurant). However, clear statements as to who was the infector between linked cases or within clusters were generally not published. As well, dates of contact between cases and details of the type of link between cases were often only reported in detail if deemed to be important for public health action, limiting the number of cases for whom detailed epidemiological information related to their linkages were available. Therefore, assumptions about directionality of transmission were largely at the discretion of the authors, and in consequence we used the following bases for identifying linked cases as directional transmission pairs: 1) linkage of the infector (but not infectee) to a cluster; 2) the dates of contact, type of contact, and onset dates reported for linked cases provided some insight into directionality of transmission; 3) the infector or index case of a chain travelled to a location with increased/increasing transmission prior to onset; or 4) the infector or index case of a chain was presumed to have been infected while travelling abroad. Households with >2 cases and links between cases in clusters where directionality of transmission and timings of contact could not be clearly identified were not selected.^55^

We included pairs with infectors who had multiple possible exposures (their exposure period takes the lower and upper bounds of all possible exposures) but excluded possible pairs where potential infectees had multiple possible infectors, and it is possible that infectees with multiple potential infectors would have different contact patterns (and possibly be associated with shorter generation intervals) compared to infectees that had only one potential infector identified, as a susceptible person is likely to become infected more quickly if they are surrounded by multiple possible sources of infection.^17,27^ Further details regarding ascertainment of transmission pairs are available in the supplementary information.

Exposures were defined in relation to travel, contact with a confirmed case, or link to a cluster/common exposure. Reports of symptom onset in Japan were not restricted to any particular symptom, such as fever, but may have been reported as beginning with any of a variety of symptoms associated with SARS-CoV-2 infection such as fever, cough, fatigue, or runny nose.

### Data stratification

The dataset including coarsely reported dates of exposure and contact were divided into strata to assess whether the generation interval, incubation period, or correlation between the two parameters would vary by subpopulation. Age (reported in deciles) was divided into three groups: cases under 30 years of age, cases 30–59 years of age, and cases 60+ years of age. Sex was reported as female or male. Separate age and sex strata were established for infectors and infectees. Type of contact between infector and infectee was divided into three categories: household contact, social contact-based interaction, and core community interaction. These divisions were made with public health interventions in mind. For example, social contact-based interaction includes types of contact that may not have occurred when local control measures were advised or a state of emergency was declared.^44^

Generally, public health control measures in Japan promoted during 2020 focused on reducing the number of people individuals were physically in contact with in a given day, as well as reducing scenarios where the “Three C’s”— closed spaces, crowded places, and close-contact settings—were present.^38^ Interventions in Japan included limiting the total number or proportion of people who can visit facilities and venues, limiting restaurant hours, encouraging staying at home and discouraging cross-prefecture travel, etc. Our definition of ore community interaction, in contrast, focuses more on contact that occurs in schools, workplaces for general business, essential workplaces (medical facilities, care facilities, government services, etc.), and unknown sources of infection (community infection). Although these settings assigned to the core community interaction category may also be targeted by public health measures, they are perhaps less acutely affected by government decrees and social sentiment compared to settings more closely related to social contact-based interaction.^44^

Japan experienced three waves of COVID-19 during 2020, with the third wave extending into 2021 (Extended Data Fig. 5). The first wave began with the first reported case, confirmed to be positive for SARS-CoV-2 on 16 January 2020. The second wave we set to begin on 1 June, which is around the center of the bottom of the trough between the peaks of the first and second waves. The third wave we set to begin on 1 October, which likewise is around the center of the bottom of the trough between the peaks of the second and third waves. Assignment to a given wave for each pair was determined by infector report date. Lastly, to check for differences given our basis for selecting transmission pairs we also stratified the dataset according to whether directionality was determined with respect to 1) importation from abroad, 2) linkage of the infector (but not infectee) to a cluster, 3) domestic travel by the infector to a location with increasing transmission, or 4) the timing and type of contact between cases in transmission chains.

### Statistical analyses

Descriptive analyses and visualization were performed using R 4.1.0.^56^ Bayesian parameter estimation was implemented in Stan using the cmdstanr interface to CmdStan 2.26.1.^57^ Data and code are available at https://github.com/nlinton/covid19_generationinterval.

To assess correlation between the generation interval and incubation period of the infector we constructed a joint probability distribution for the generation interval and infector incubation period by use of copulas (multivariate cumulative distribution functions).^58,59^ The copulas we assessed included the Gaussian (normal), Clayton, Gumbel, and independence copulas. They are described in detail in the Supplementary Materials. Timing of pathogen transmission and symptom onset was estimated using interval censoring methods derived from Reich et al.^60^ and adapted from previously published work.^1,5^ For all parameters, posterior point estimates are given by the 50th percentiles of the converged Markov chain Monte Carlo (MCMC) chains from 100,000 iterations, and the best combination of copula and parametric distributions were selected using weights from a Bayesian mixture model (see Supplementary Materials).

To consider the effect of correlation between the generation interval and incubation period on transmission, we simulated 10,000 pairs from the best-fit model of the jointly estimated generation interval and incubation period for all possible values of Kendall’s tau from 0 to 1. For each simulated pair, we determined whether transmission was presymptomatic based on whether the incubation period was greater than the generation interval, and thereby calculated the proportion of presymptomatic transmission *p* for the 10,000 pairs for each value of Kendall’s tau. We then considered that symptomatic transmission could result in a decrease in transmission as defined by the basic reproduction number *R*_0_—the average number of infectees generated by a single infector. We calculated the effective *R*_0_ as 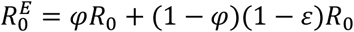 where *φ* is the proportion of presymptomatic transmission and *ε* is the percent reduction in transmission due to rapid isolation among symptomatic cases, considering *R*_0_ = 2.2^4,30^ and varied this by ±0.7, also considering *R*_0_ = 1.5 and *R*_0_ = 2.9.

## Results

### Characteristics of transmission pairs

Information on timing of exposure and onset for infectors as well as contact between infectors and infectees was obtained for 286 transmission pairs of confirmed cases reported in Japan during 2020, of which 257 pairs had symptom onset available for the infector. For the other 29 pairs the infectors were asymptomatic at time of report. Of the 257 pairs with symptomatic infectors, 49 (19.2%) had single dates reported for both infector exposure and contact between infector and infectee. Characteristics of the pairs in each dataset are shown in Table 1, while Extended Data Fig. 1 provides insight into the relationship between the empirical generation intervals, serial intervals, and incubation periods associated with these cases.

**Table 1.** Characteristics of COVID-19 transmission pairs in Japan, 2020

| Strata | Role | Characteristics | Coarse dates of exposure<sup>†</sup> (%) | Single dates of exposure<sup>‡</sup> (%) | Asymptomatic infectors (%) |
| --- | --- | --- | --- | --- | --- |
|  |  | All | 257 pairs | 49 pairs | 29 pairs |
| Age | Infector | Under 30 years | 65 (25.3%) | 11 (22.4%) | 6 (20.7%) |
|  |  | 30-59 years | 129 (50.2%) | 32 (65.3%) | 15 (51.7%) |
|  |  | 60+ years | 57 (22.2%) | 6 (12.2%) | 7 (24.1%) |
|  |  | Not reported | 6 (2.3%) | - | 1 (3.4%) |
|  | Infectee | Under 30 years | 69 (26.8%) | 17 (34.7%) | 9 (31.0%) |
|  |  | 30-59 years | 114 (44.4%) | 19 (38.8%) | 10 (34.5%) |
|  |  | 60+ years | 67 (26.1%) | 13 (26.5%) | 9 (31.0%) |
|  |  | Not reported | 7 (2.7%) | - | 1 (3.4%) |
| Sex | Infector | Female | 79 (30.7%) | 17 (34.7%) | 12 (41.4%) |
|  |  | Male | 177 (68.9%) | 32 (65.3%) | 16 (55.2%) |
|  |  | Not reported | 1 (0.4%) | - | 1 (3.4%) |
|  | Infectee | Female | 131 (51.0%) | 22 (44.9%) | 16 (55.2%) |
|  |  | Male | 125 (48.6%) | 27 (55.1%) | 12 (41.4%) |
|  |  | Not reported | 1 (0.4%) | - | 1 (3.4%) |
| Transmission setting | Infector | Household | 4 (1.6%) | 0 (0.0%) | 0 (0.0%) |
|  |  | Social | 153 (59.5%) | 48 (98.0%) | 25 (86.2%) |
|  |  | Community | 100 (38.9%) | 1 (2.0%) | 4 (13.8%) |
|  | Infectee | Household | 109 (42.4%) | 2 (4.1%) | 11 (37.9%) |
|  |  | Social interaction | 113 (44.0%) | 38 (77.6%) | 15 (51.7%) |
|  |  | Community | 35 (13.6%) | 9 (18.4%) | 3 (10.3%) |
| Epidemic wave |  | Wave 1 | 86 (33.5%) | 14 (28.6%) | 2 (6.9%) |
|  |  | Wave 2 | 95 (37.0%) | 16 (32.7%) | 13 (44.8%) |
|  |  | Wave 3 | 76 (29.6%) | 19 (38.8%) | 14 (48.3%) |
| Basis for selection |  | Cluster | 138 (53.7%) | 29 (59.2%) | 24 (82.8%) |
|  |  | Contact pattern | 54 (21.0%) | 19 (38.8%) | 4 (13.8%) |
|  |  | Domestic travel | 43 (16.7%) | 1 (2.0%) | 1 (3.4%) |
|  |  | Import | 22 (8.6%) | - | - |
| Region§ |  | Hokkaido & Tohoku | 35 (13.6%) | 6 (12.2%) | 2 (6.9%) |
|  |  | Kanto | 31 (12.1%) | 4 (8.2%) | 0 (0.0%) |
|  |  | Chubu | 73 (28.4%) | 12 (24.5%) | 11 (37.9%) |
|  |  | Kinki | 38 (14.8%) | 6 (12.2%) | 3 (10.3%) |
|  |  | Chugoku & Shikoku | 34 (13.2%) | 6 (12.2%) | 6 (20.7%) |
|  |  | Kyushu & Okinawa | 46 (17.9%) | 15 (30.6%) | 7 (24.1%) |

<sup>†</sup>Dataset using intervals of exposure and/or contact between infector and infectee. <sup>‡</sup>Dataset limited to pairs where infector exposure and infector-infectee contact were limited to a single day. “Hokkaido & Tohoku” includes Hokkaido, Aomori, Iwate, Miyagi, Akita, Yamagata, and Fukushima prefectures. “Kanto” includes Ibaraki, Tochigi, Gunma, Saitama, Chiba, Tokyo, and Kanagawa prefectures. “Chubu” includes Niigata, Toyama, Ishikawa, Fukui, Yamanashi, Nagano, Gifu, Shizuoka, and Aichi prefectures. “Kinki” includes Mie, Shiga, Kyoto, Osaka, Hyogo, Nara, and Wakayama prefectures. “Chugoku & Shikoku” include Tottori, Shimane, Hiroshima, Yamaguchi, Tokushima, Kagawa, Ehime, and Kochi prefectures. “Kyushu & Okinawa” include Fukuoka, Saga, Nagasaki, Kumamoto, Oita, Miyazaki, Kagoshima, and Okinawa prefectures.

For the dataset of 257 pairs with symptomatic infectors, most infectors (50.2%) and infectees (44.4%) were between 30–59 years of age. There were fewer female infectors (30.7%) detected compared to female infectees (51.0%). Age and sex distributions of infectors and infectees are shown in Extended Data Fig. 2. Pairs were relatively evenly distributed between the three pandemic waves that occurred during 2020. Most pairs (53.7%) were linked to a cluster—an aggregation of cases with a common exposure, while other pairs were identified by having contact patterns indicative of directionality of transmission (21.0%), the infector had travel to another prefecture before onset (16.7%), or the infector was an imported case or otherwise linked to an imported case (8.6%). Given that it was easier to determine the directionality of pairs and obtain information on timing of exposure if the infector was linked to a cluster or had travel history, our dataset includes only a handful of infectors (1.6%) with household exposure. In contrast, nearly half (42.4%) of infectees were household/family members of their infectors. The single-date (49 pairs) and asymptomatic infector (28 pairs) datasets were similarly structured in terms of age and sex, though only one asymptomatic infector was detected during the first wave.

### Joint estimates of the generation interval and incubation period

The jointly estimated mean generation interval ranged between 3.7 and 5.1 days, with the mean for the overall dataset estimated at 4.3 days (95% CrI: 4.0–4.7 days). In contrast, the estimated generation interval for the dataset of asymptomatic infectors was longer, at 4.6 days (3.9–5.5 days), resulting in a ratio of asymptomatic-to-symptomatic generation intervals of 1.1 The jointly estimated mean incubation period was consistently longer than the generation interval, ranging from 4.4–5.7 days, and estimated at 4.8 days (95% CrI: 4.4–5.1 days) for the overall dataset, providing evidence of presymptomatic transmission. The prior and posterior distributions of the generation interval and incubation period are shown in Extended Data Fig. 3. The generation interval and incubation period were positively correlated, with Kendall’s tau ranging between 0.4–0.6 and estimated at 0.5 (95% CrI: 0.4–0.6) for the overall dataset (Table 2). For the dataset with single dates of reported exposure and contact, the generation interval was estimated at 4.4 days (95% CrI: 3.9–5.0 days) while the mean incubation period was estimated at 4.9 days (95% CrI: 4.4–5.6 days). Kendall’s tau was slightly higher than for the overall dataset, at 0.6 (95% CrI: 0.5–0.7).

**Table 2.** Joint estimates of the generation interval and incubation period of COVID-19 cases by stratum

| Category | Strata | N | Copula | Generation interval |  |  | Incubation period |  |  | Kendall's tau<br>(95% CrI) |
| --- | --- | --- | --- | --- | --- | --- | --- | --- | --- | --- |
|  |  |  |  | Distribution | Mean (95% CrI) | SD (95% CrI) | Distribution | Mean (95% CrI) | SD (95% CrI) |  |
| Exact data | All cases | 49 | Clayton | Weibull | 4.38 (3.88, 4.98) | 2.10 (1.70, 2.79) | Lognormal | 4.91 (4.34, 5.60) | 2.66 (2.18, 3.33) | 0.61 (0.46, 0.68) |
| Coarse data | All cases | 257 | Clayton | Weibull | 4.34 (3.98, 4.74) | 2.31 (2.00, 2.81) | Lognormal | 4.75 (4.42, 5.11) | 2.57 (2.25, 2.98) | 0.54 (0.44, 0.63) |
| Infector<br>age | Under 30 years | 65 | Clayton | Lognormal | 4.51 (3.95, 5.13) | 2.15 (1.68, 2.89) | Lognormal | 4.67 (4.06, 5.36) | 2.60 (2.12, 3.27) | 0.50 (0.23, 0.65) |
|  | 30-59 years | 129 | Gumbel | Gamma | 4.43 (3.97, 4.94) | 2.43 (2.01, 3.00) | Lognormal | 5.10 (4.63, 5.62) | 2.87 (2.45, 3.44) | 0.60 (0.48, 0.68) |
|  | 60+ years | 57 | Gumbel | Gamma | 4.58 (3.87, 5.37) | 2.68 (1.98, 3.68) | Lognormal | 4.59 (3.91, 5.37) | 2.89 (2.31, 3.74) | 0.50 (0.28, 0.65) |
| Infected<br>age | Under 30 years | 69 | Clayton | Gamma | 4.57 (4.02, 5.20) | 2.39 (1.90, 3.18) | Lognormal | 4.72 (4.15, 5.38) | 2.62 (2.15, 3.26) | 0.56 (0.37, 0.68) |
|  | 30-59 years | 114 | Gumbel | Weibull | 4.66 (4.13, 5.22) | 2.59 (2.07, 3.25) | Lognormal | 5.26 (4.72, 5.85) | 3.12 (2.62, 3.78) | 0.58 (0.47, 0.68) |
|  | 60+ years | 67 | Gumbel | Lognormal | 4.10 (3.48, 4.83) | 2.08 (1.49, 3.01) | Lognormal | 4.45 (3.84, 5.15) | 2.59 (2.10, 3.32) | 0.42 (0.14, 0.61) |
| Infector sex | Female | 79 | Gumbel | Gamma | 3.72 (3.20, 4.35) | 2.09 (1.57, 2.92) | Lognormal | 4.38 (3.82, 5.02) | 2.59 (2.10, 3.26) | 0.54 (0.36, 0.66) |
|  | Male | 177 | Clayton | Weibull | 4.71 (4.29, 5.20) | 2.36 (2.00, 2.90) | Lognormal | 5.00 (4.60, 5.46) | 2.73 (2.35, 3.22) | 0.52 (0.39, 0.63) |
| Infected sex | Female | 131 | Clayton | Gamma | 4.35 (3.82, 4.95) | 2.50 (1.99, 3.28) | Lognormal | 4.65 (4.20, 5.16) | 2.60 (2.20, 3.15) | 0.50 (0.33, 0.64) |
|  | Male | 125 | Gumbel | Gamma | 4.50 (4.05, 4.99) | 2.28 (1.88, 2.87) | Lognormal | 4.96 (4.48, 5.52) | 2.88 (2.43, 3.48) | 0.55 (0.43, 0.65) |
| Epidemic<br>wave | Wave 1 | 109 | Gumbel | Weibull | 4.38 (3.74, 5.01) | 1.99 (1.59, 2.58) | Lognormal | 4.75 (4.21, 5.36) | 2.93 (2.46, 3.63) | 0.53 (0.31, 0.67) |
|  | Wave 2 | 113 | Clayton | Gamma | 4.50 (4.04, 5.03) | 2.50 (2.09, 3.13) | Lognormal | 4.85 (4.39, 5.38) | 2.55 (2.16, 3.07) | 0.58 (0.47, 0.67) |
|  | Wave 3 | 35 | Gaussian | Gamma | 4.52 (3.81, 5.32) | 2.39 (1.74, 3.43) | Lognormal | 4.98 (4.19, 5.87) | 3.01 (2.38, 3.93) | 0.57 (0.36, 0.68) |
| Transmission<br>setting | Household | 86 | Gumbel | Weibull | 5.09 (4.46, 5.77) | 2.82 (2.25, 3.59) | Lognormal | 5.13 (4.53, 5.79) | 2.87 (2.38, 3.55) | 0.55 (0.38, 0.67) |
|  | Social contact | 95 | Gumbel | Weibull | 3.84 (3.42, 4.33) | 1.74 (1.38, 2.32) | Lognormal | 4.53 (4.02, 5.12) | 2.68 (2.22, 3.34) | 0.55 (0.40, 0.66) |
|  | Community | 76 | Clayton | Gamma | 4.45 (3.87, 5.11) | 2.36 (1.85, 3.20) | Lognormal | 5.00 (4.40, 5.69) | 2.93 (2.42, 3.68) | 0.48 (0.29, 0.63) |
| Basis for<br>selection | Cluster | 138 | Gumbel | Gamma | 4.08 (3.62, 4.60) | 2.17 (1.73, 2.81) | Lognormal | 4.55 (4.14, 5.02) | 2.50 (2.13, 2.99) | 0.46 (0.29, 0.60) |
|  | Contact pattern | 54 | Clayton | Weibull | 4.62 (4.03, 5.30) | 2.31 (1.82, 3.11) | Lognormal | 5.12 (4.45, 5.90) | 3.07 (2.48, 3.92) | 0.59 (0.42, 0.68) |
|  | Domestic travel | 43 | Clayton | Gamma | 4.84 (4.12, 5.64) | 2.64 (2.01, 3.62) | Lognormal | 5.01 (4.22, 5.92) | 3.02 (2.40, 3.93) | 0.47 (0.23, 0.64) |
|  | Import | 22 | Gaussian | Weibull | 4.83 (3.91, 5.78) | 2.33 (1.62, 3.50) | Lognormal | 5.65 (4.60, 6.77) | 3.49 (2.73, 4.55) | 0.64 (0.37, 0.69) |
| Asymptomatic infectors |  | 29 | - | Gamma | 4.62 (3.78, 5.61) | 2.45 (1.78, 3.59) | - | - | - | - |

CrI: credible interval. Wave 1 began 16 January 2020. Wave 2 is assumed to have begun 1 June 2020, and wave 3 is assumed to have begun 1 October 2020.

The mean generation interval did not vary substantially between strata but was shortest for female infectors, at 3.7 days (95% CrI: 3.2–4.4 days). It was also shorter for the second wave of the epidemic (3.8 days, 95% CrI: 3.4–4.3 days) compared to the first wave (5.1 days, 95% CrI: 4.5–5.8 days).^27^ However, the generation interval for the third wave was longer than that of the second wave—nearly as long as that of the first wave—at 4.5 days (95% CrI: 3.9–5.1 days). Estimates of the incubation period varied less between strata, although the mean incubation period for pairs linked to importation from other countries (mostly from the first wave) was a bit longer than the overall estimate, at 5.7 days (95% CrI: 4.6–6.8 days).

The Clayton copula—which emphasizes lower tail dependence—was the most frequently selected copula, although the Gumbel and Gaussian copulas were also selected for some strata. The Gumbel copula emphasizes upper tail dependence while the Gaussian copula does not consider tail dependence. The independence copula was never selected (Table 2). For the overall dataset, where the Clayton copula was selected, the lower tail dependence was 0.7 (95% CrI: 0.6–0.8), indicating that infectors with an extremely short incubation period would also be more likely to quickly transmit the virus given contact with a susceptible person (see Supplementary Materials). For the generation interval, the Weibull distribution was most often selected, although the gamma and lognormal distributions were selected for some strata. The lognormal distribution was the only distribution selected across all joint estimates of the incubation period. It is typically the best fit for infectious disease incubation period data,^28^ including COVID-19 data.^29^

### Correlation, presymptomatic transmission, and control measures

We found that 63.2% of pairs experienced presymptomatic transmission, defined as the generation interval being shorter than the incubation period, by simulating 10,000 transmission pairs from our fitted estimates of the generation interval, incubation period, and Kendall’s tau (Table 2). Using this fit, we varied Kendall’s tau for the same estimates of the generation interval and incubation period and show that as Kendall’s tau approached zero (independence) the proportion of presymptomatic transmission reached a lower boundary of 54.2%. Conversely, as Kendall’s tau approached 1 (complete dependence), the proportion of presymptomatic transmission increased to nearly 100% (Figure 3a), and the difference between symptom onset and transmission became so small that they mostly occurred on the same day, with only a small portion of presymptomatic transmission occurring outside of one day before or after symptom onset, and no symptomatic transmission occurring.

**Figure 1.**
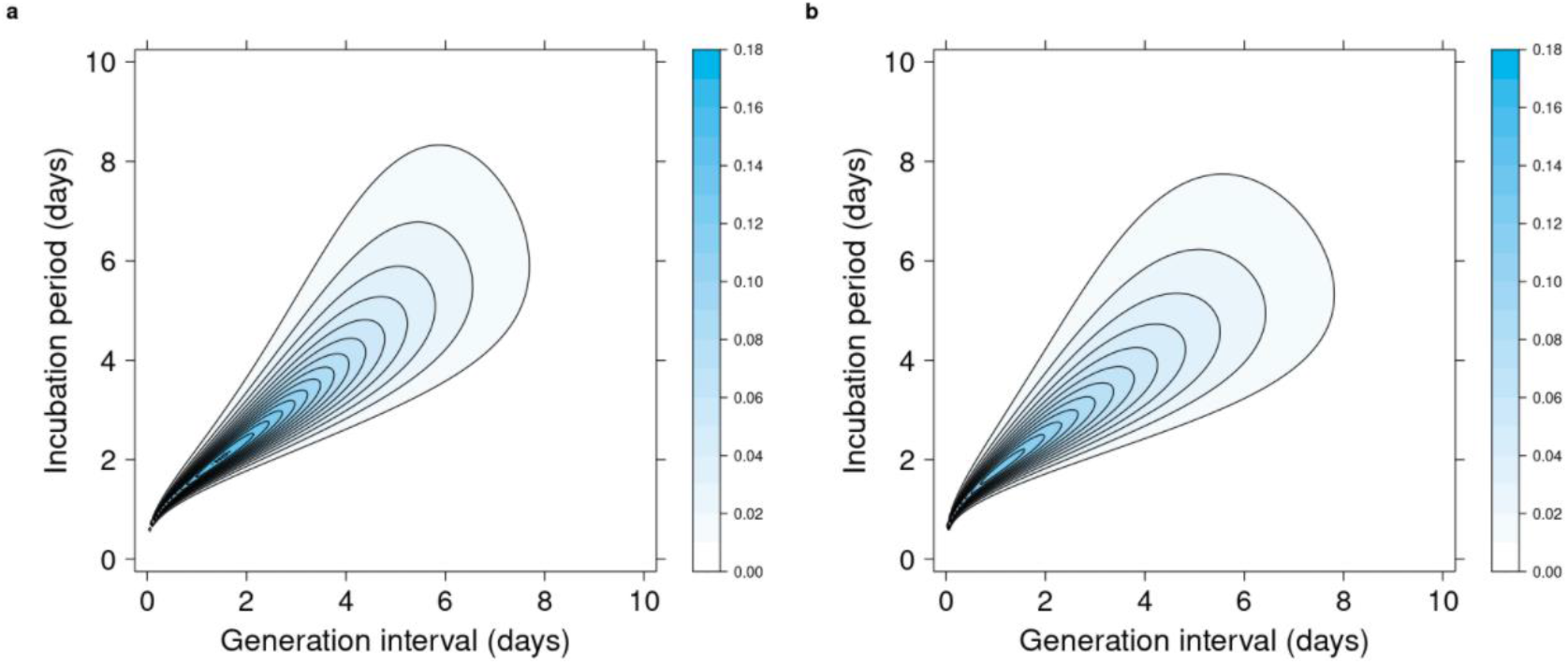
Joint distribution of the generation interval and incubation period. Contour plots of the fitted distributions. For both a, the dataset of 49 transmission pairs with single dates of reported exposure, and b, the dataset of 257 transmission pairs that also includes pairs with more coarsely reported possible dates of exposure and contact, a Clayton copula with a Weibull marginal for the generation interval and lognormal marginal for the incubation period distribution was selected.

**Figure 2.**
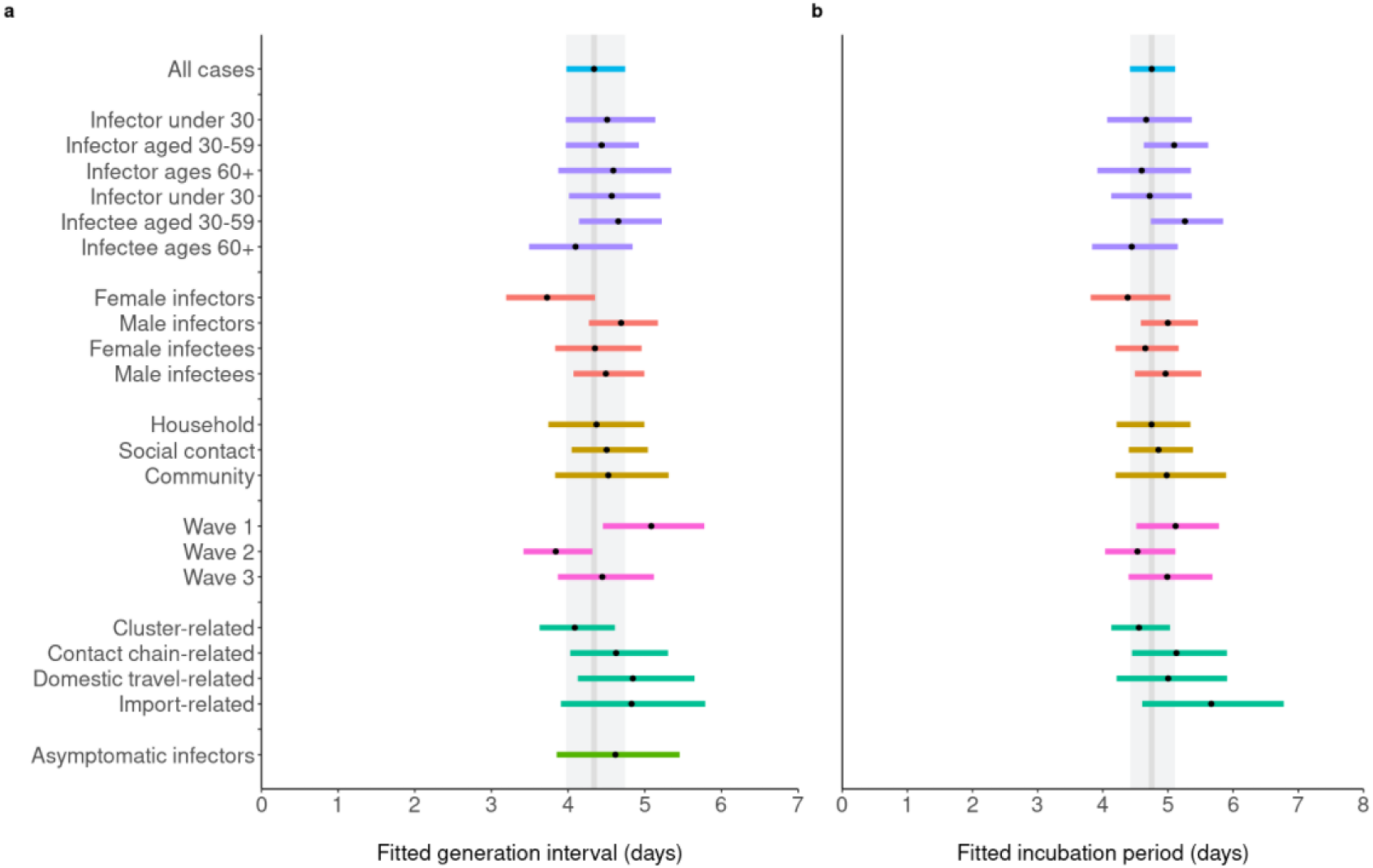
Joint estimates of the generation interval and incubation period by stratum for COVID-19 transmission pairs from Japan. The joint distribution using best-fit Gaussian, Gumbel, or Clayton copula combined with gamma, lognormal, or Weibull distributions for the **a**, generation interval and **b**, incubation period are presented for the dataset of 257 transmission pairs that also includes pairs with more coarsely reported possible dates of exposure and contact. The points are point estimates for the means of each stratum, while the colored bars indicate the 95% credible intervals. The grey bars show the overall point estimate and 95% CrI for all cases in the background. The estimate for asymptomatic infectors was fitted to the generation interval alone, as infector incubation period could not be estimated.

**Figure 3.**
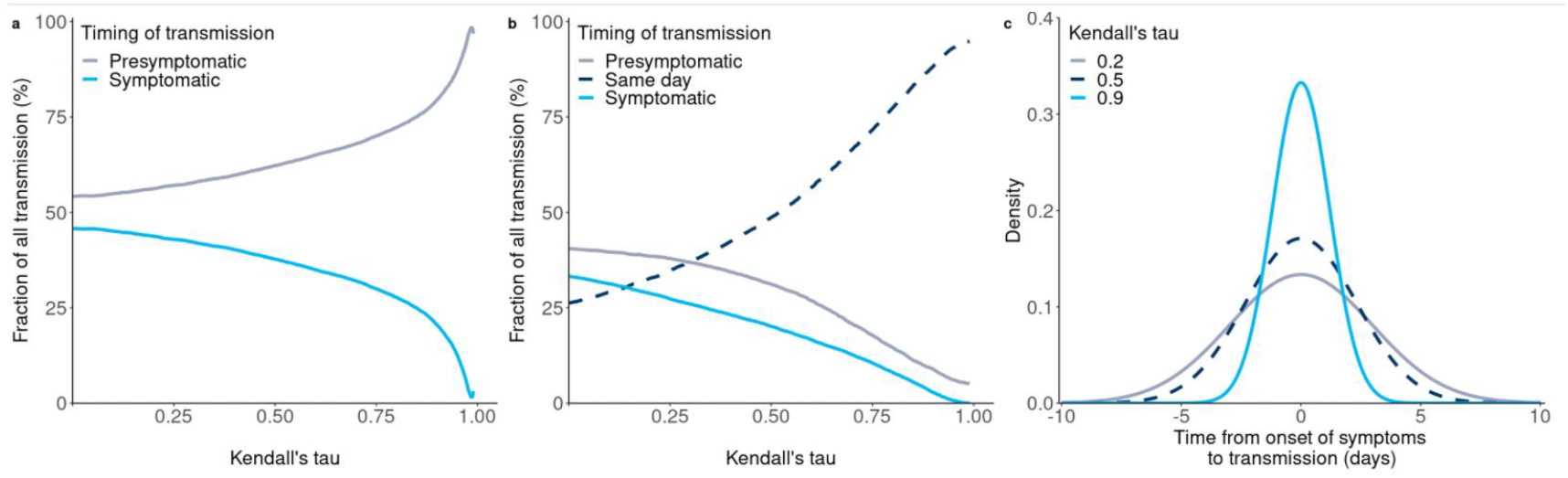
Increased correlation leads to a predominance of presymptomatic transmission. From estimates made by simulating the generation interval (GI) and incubation period (IP) for 10000 pairs using the fitted Clayton copula with Weibull (GI) and lognormal (IP) marginals: **a**, the proportion of transmission that was symptomatic or presymptomatic for various values of Kendall’s tau; **b**, the proportion of transmission that was symptomatic, presymptomatic, or occurred on the same day (GI-IP ∈ [−**1, 1**]) for various values of Kendall’s tau; c, the time from onset of symptoms to transmission (TOST), defined as GI-IP, fitted with a normal distribution.

The average difference between the generation interval and incubation period across all data subsets and strata was 0.4 days, indicating a mean time from onset of symptoms to transmission of -0.4 days. The probability density function of the time from onset of symptoms to transmission fitted with a normal distribution based on simulated data with Kendall’s tau varied between 0.2, 0.5, and 0.9, is shown in Figure 3c. The mean was centered at -0.4 days, and lower correlation resulted in a larger standard deviation.

Using the same simulated dataset, Kendall’s tau was varied against the fraction of transmission reduced by case isolation. Increasing Kendall’s tau indicated greater difficulty in controlling transmission via isolation alone given the same level of reduction in transmission due to rapid isolation *φ* (Figure 4). Given a basic reproduction number of *R*_0_ of 2.2 or 2.9,^30^ the effective *R*_0_ (denoted 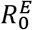) failed to reduce below the epidemic threshold of 1 (Figure 4b and c). Moreover, as Kendall’s tau approaches 1 and the proportion of presymptomatic transmission (with a mean time from onset of symptoms to transmission of approximately 0) approaches 100%, control through isolation alone becomes impossible, and there is no difference between *R*_0_ and 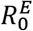. When case isolation does not occurs promptly following onset (lower *φ*) the dependence between the generation interval and incubation period has less impact on the reduction of 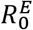.

**Figure 4.**
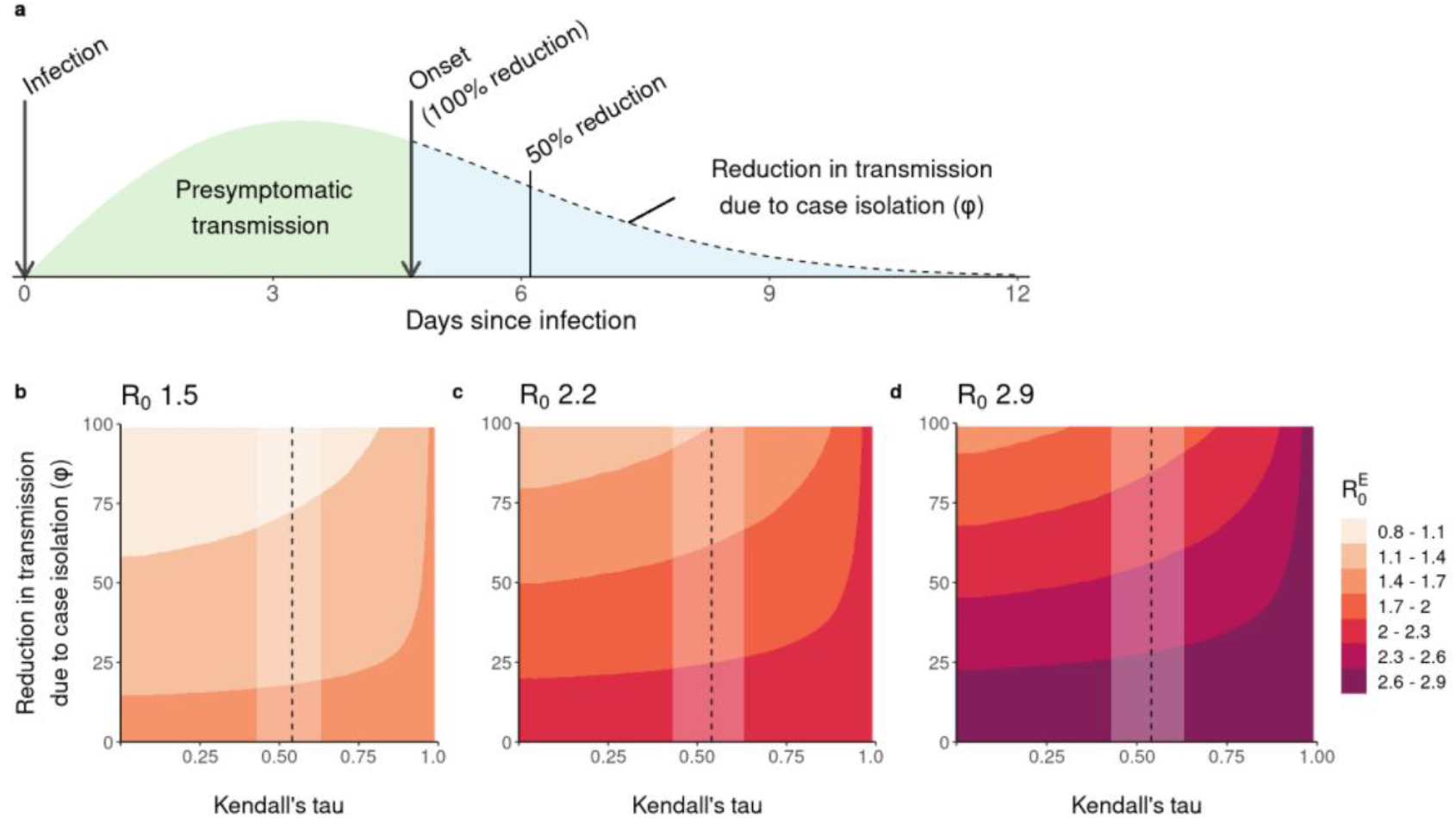
Effect of stronger correlation between the generation interval and incubation period on effectiveness of isolation. The top figure, **a**, shows the fitted generation interval probability distribution function (4.22 days). The arrow dividing the green and blue sections indicates onset at 4.67 days (the mean incubation period). If case isolation occurs at onset this is equivalent to a 100% reduction in possible transmission for symptomatic cases. Figures **b, c**, and **d**, show the effective basic reproduction number 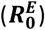 as a function of this reduction in transmission as well as the level of correlation between the generation interval and incubation period. The dashed line is the point estimate of Kendall’s tau obtained in this study, while the shaded white rectangle shows its 95% credible interval. We assume baseline ***R***_**0**_ of **b**, 1.5, **c**, 2.2, and **d**, 2.9. As the generation interval and incubation period approach independence (Kendall’s tau→ **1**) case isolation will become ineffective—shown by the unchanging effective 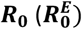— as transmission will either be presymptomatic or occur nearly at the same time as symptom onset.

## Discussion

The generation interval underpins many infectious disease models,^31,32^ and here we provide insight into the generation interval of COVID-19 over time and across different characteristics of transmission pairs, providing one of the most comprehensive characterizations of the generation interval of wild-type COVID-19 to date. In addition, we quantitatively measured the relationship between the generation interval and incubation period of COVID-19, the lack of which was identified as a limiting factor in previous studies.^7,23,33^ From transmission pairs identified using publicly available data reported in Japan during 2020 we found positive correlation between the generation interval and incubation period with a Kendall’s tau ranging from 0.4–0.6. The mean generation interval was consistently shorter than the mean incubation period when jointly estimated, with the former ranging between 3.7–5.1 days and the latter between 4.4–5.7 days, indicating consistent presence of presymptomatic transmission.

The means of the jointly estimated generation interval and incubation period are in line with those reported elsewhere^6–8,10^ with the mean generation interval reported here—4.3 days—falling in the range of 2.8–7.5 days previously reported (see Supplementary Table 1 and Extended Data Fig. 4). The positive correlation between the generation interval and incubation period indicates that for symptomatic cases, onset is tied to infectiousness. This finding supports evidence shown in virological studies.^25^ The estimate of the generation interval estimated with asymptomatic infectors (4.6 days, 95%: 3.8, 5.6 days) was longer than the jointly estimated generation interval using symptomatic infectors indicating that infectiousness in asymptomatic cases may be more persistent than in symptomatic infections—potentially leading to an underestimation of *R*_0_ using estimates from symptomatic pairs.^34^ Although evidence has indicated that asymptomatic COVID-19 cases are less infectious than symptomatic cases,^20^ asymptomatic cases nonetheless play a notable role in epidemic dynamics.^6^

The proportion of presymptomatic transmission among symptomatic cases estimated in this study, 63.2%, is higher than estimates reported by published studies using data from early in the pandemic,^6,25^ but is similar to other estimates.^7,8,35^ Among those pairs with presymptomatic transmission, 33.5% had transmission occurring within one day of infector onset, and 49.1% within two days of infector onset. Combining estimates of asymptomatic transmission (in the range of 18–30%)^36,37^ with the estimate of presymptomatic transmission shared here, the proportion of nonsymptomatic transmission could feasibly reach 90%. Thus, interventions such as physical distancing that do not depend on detection of potential infectors while they are not showing symptoms, enhanced surveillance to detect nonsymptomatic cases, and contact tracing to identify exposed individuals while their infected contacts are not symptomatic are crucial for COVID-19 control.^6,18,19^

Data collection for this study focused on high certainty of directionality of transmission based on publicly announced epidemiological data. In contrast to most other countries, Japan applied backward contact tracing methods from the beginning of the pandemic in an effort to prevent large clusters of cases,^38^ making it an apt setting for obtaining transmission pair data. This is because links between cases—and particularly those related to clusters—were more likely to have been detected compared to countries where backward contact tracing was not conducted. However, the timing of COVID-19 testing plays an important role in case ascertainment,^39^ and infected persons who were epidemiologically linked to COVID-19 cases but did not become symptomatic after initially testing negative for SARS-CoV-2 may have been missed as cases. In addition, Japan did not promote widespread community viral testing, and this perhaps limited the number of unlinked cases that may have otherwise been detected and retrospectively linked to others during the epidemic. As well, public health jurisdiction reporting practices changed over time, with details generally becoming sparser once daily incidence became high enough to wear contact tracing capacity thin, and also towards the end of the year as concerns grew around infection-related stigmatization.^40^

The shorter mean generation interval during the second and third waves of pandemic in Japan compared to the first wave may in part reflect the increase in prevalence of infection, as increased competition between infectious individuals to find susceptible contacts can lead to contraction of the generation interval.^17,27,41^ Shorter generation intervals were also noted in the United Kingdom during September– November 2020, when there was a rise in the number of new cases.^35^ However, our results indicate that increases in incidence do not perforce lead to contraction of the generation interval. The larger value obtained for the third wave in Japan, which had a higher peak than the second wave (Extended Data Fig. 5).

During the first epidemic wave, state of emergency declarations nationwide. However, during the second wave and the 2020 half of the third wave, no such preventative measures were introduced. Conversely, campaigns intending to restart the Japanese economy following the difficulties caused by the first wave of COVID-19 were developed and implemented. In particular, the GoTo Travel campaign, which offered discounts on travel inside Japan, was a fixture of the second and third waves (Extended Data Fig. 5). The campaign began just before the peak of the second wave and was associated with an increase in COVID-19 cases reporting inter-prefecture travel.^42^ Of our pairs identified for the second wave, only 20.0% of infectors were reported before the start of the GoTo travel campaign. Although we did not find that our pairs where the infector had domestic travel experienced longer generation intervals (Figure 2), travel can left-censor the time following infection when an infector who travelled had contact with an infectee who did not travel with them.

Most identified transmission pairs had contact in the household or in settings related to social behavior, such as eating at restaurants, visiting nightlife, singing karaoke, attending sports events, listening to live music, visiting gyms, or meeting with friends, relatives, acquaintances, etc. (Table 1). These types of social contact settings have also been associated with SARS-CoV-2 transmission in other countries.^43^ In Japan, settings for social contact first are the first to be requested to be restricted by prefectural and local governments when control measures or a state of emergency was deemed necessary to reduce case incidence,^44^ though such emergency measures were not implemented during the second or third wave portions of 2020. Similar interventions focused on limiting social contact were implemented in other parts of the world,^45^ though other countries had a greater focus on reducing formalized community contact, such as by moving schools and workplaces online.^46,47^

As SARS-CoV-2 variants begin to dominate transmission in many countries,^48–50^ it remains to be seen whether the mean and variance of the generation interval and incubation period for the new variants of concern (VOC) will be similar to estimates presented in this, or previous studies. It has been suggested that the Alpha (Pango lineage B.1.1.7) variant could have longer generation intervals,^51^ and may therefore be more responsive to interventions targeted towards speed, such as contact tracing.^52^ However, a study using Singapore transmission pairs did not find a large difference (>1 day) between the serial interval of wild-type and Delta (Pango lineage B.1.617.2) variant SARS-CoV-2 infections.^53^ As the mean serial interval can approximate the mean generation interval, this finding may indicate that the generation interval of the Delta variant will not vary much from that of wild-type SARS-CoV-2. Similarly, a preprint analysis of Delta variant cases from China estimated an incubation period of 5.8 days,^54^ which resembles the results from a meta-analysis on wild-type SARS-CoV-2 incubation periods.^29^

Whether the correlation between the generation interval and incubation period would be weaker or stronger than has been presented here remains to be seen. However, this study provides detailed estimates of the generation interval, incubation period, and their correlation for different groups and time periods in Japan. In doing so, it provides basis for consideration of correlation going forward.

## Data Availability

The data and code for this study are available on GitHub at https://github.com/nlinton/covid19_generationinterval

## Extended Data figures

**Extended Data Fig. 1.**
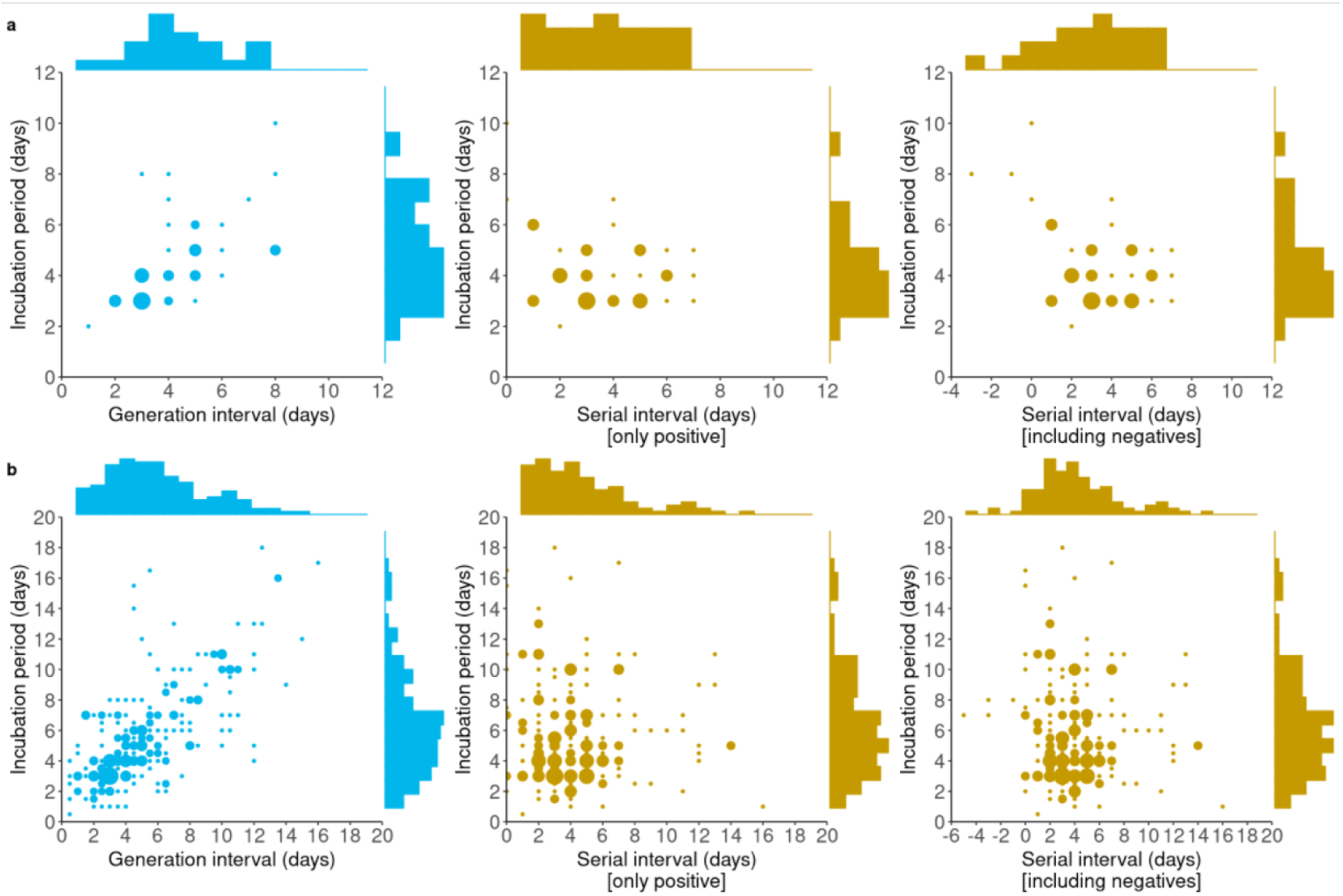
Correlation of empirical generation and serial intervals with the incubation period of COVID-19 cases in Japan. Scatterplots showing the generation and serial intervals plotted against the incubation period for transmission pairs with single-date (n=49) and coarsely observed (n=257) exposures. For **a**, data where single dates of exposure for the infector and contact between infector and infectee were reported, Kendall’s tau was 0.58 (p<0.001), -0.13 (p=0.32), and -0.23 (p=0.07) for the generation interval, serial interval with only positives, and serial interval including negatives, respectively. For **b**, data with coarse dates of exposure and contact, the plotted value represents the midpoint of the possible exposure/contact period, and Kendall’s tau was 0.53 (p<0.001), -0.02 (p=0.66), and -0.04 (p=0.39).

**Extended Data Fig. 2.**
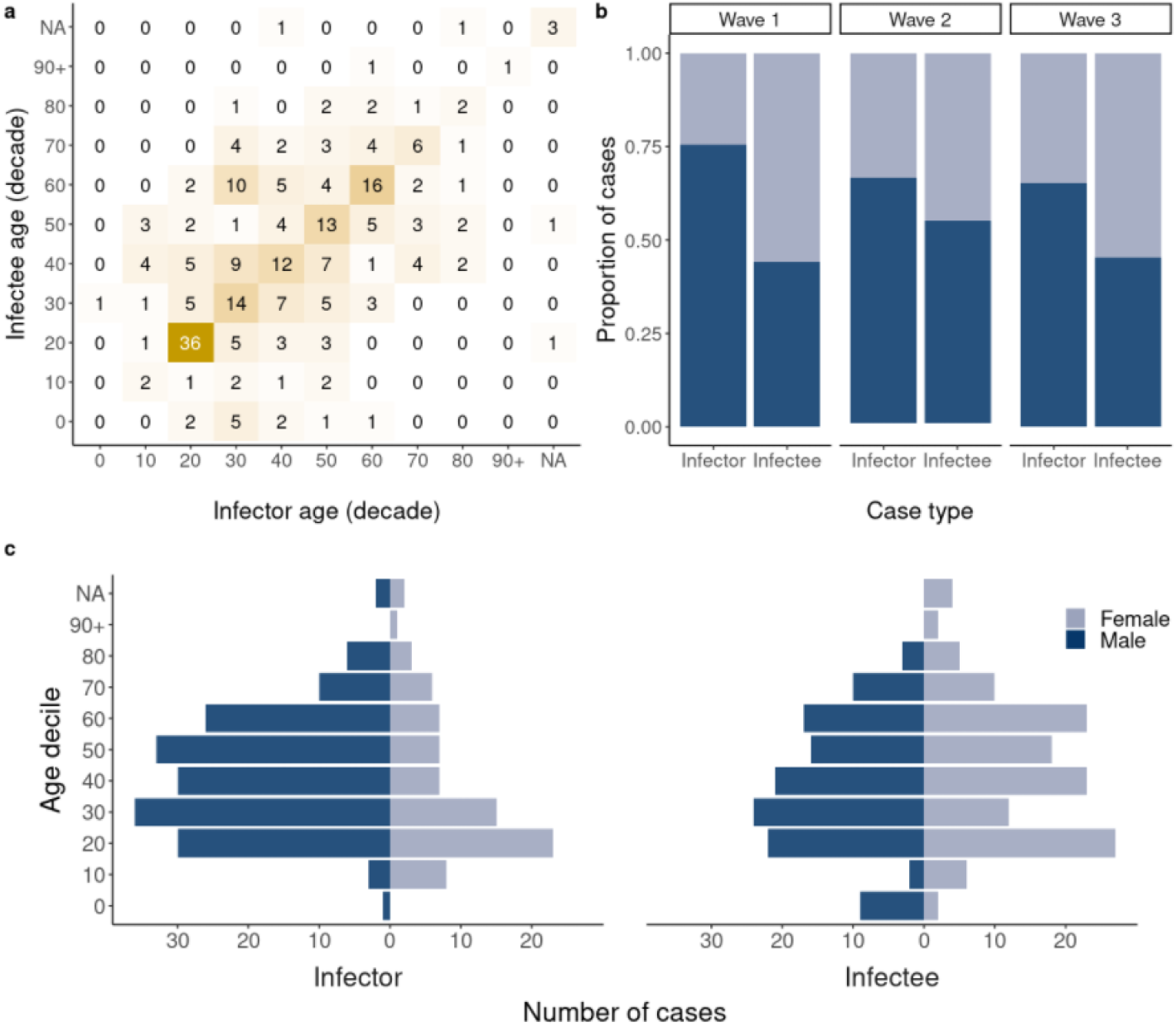
Age and sex distribution of transmission pairs. **a**, Matrix of infector and infectee age groups for 257 transmission pairs. **b**, Proportion of cases by sex, wave, and infector-infectee status. **c**, Age group and sex distribution of infectors and infectees.

**Extended Data Fig. 3.**
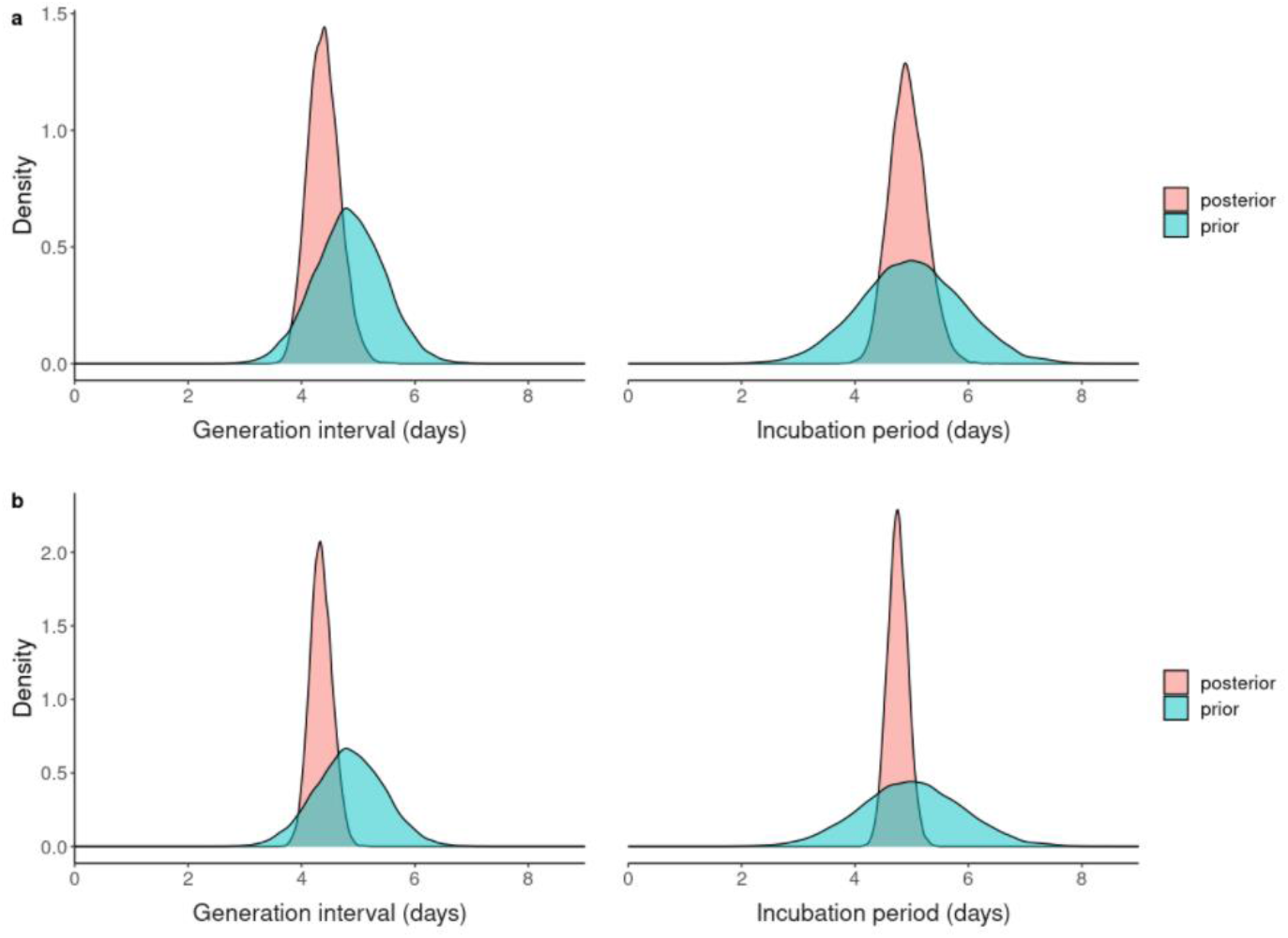
Prior and posterior densities of the mean. **a**, densities for the dataset where single dates of exposure for the infector and contact between infector and infectee were reported (n=49). **b** densities for the dataset with coarsely reported dates of exposure and contact (n=257). The priors are the same for **a** and **b**; for the generation interval the prior was obtained from the values of the serial interval reported by Nishiura et al., while the prior for the incubation period was obtained from the values of the incubation period reported by Linton et al.—both studies used cases reported worldwide during the early stages of the pandemic (early 2020).

**Extended Data Fig. 4.**
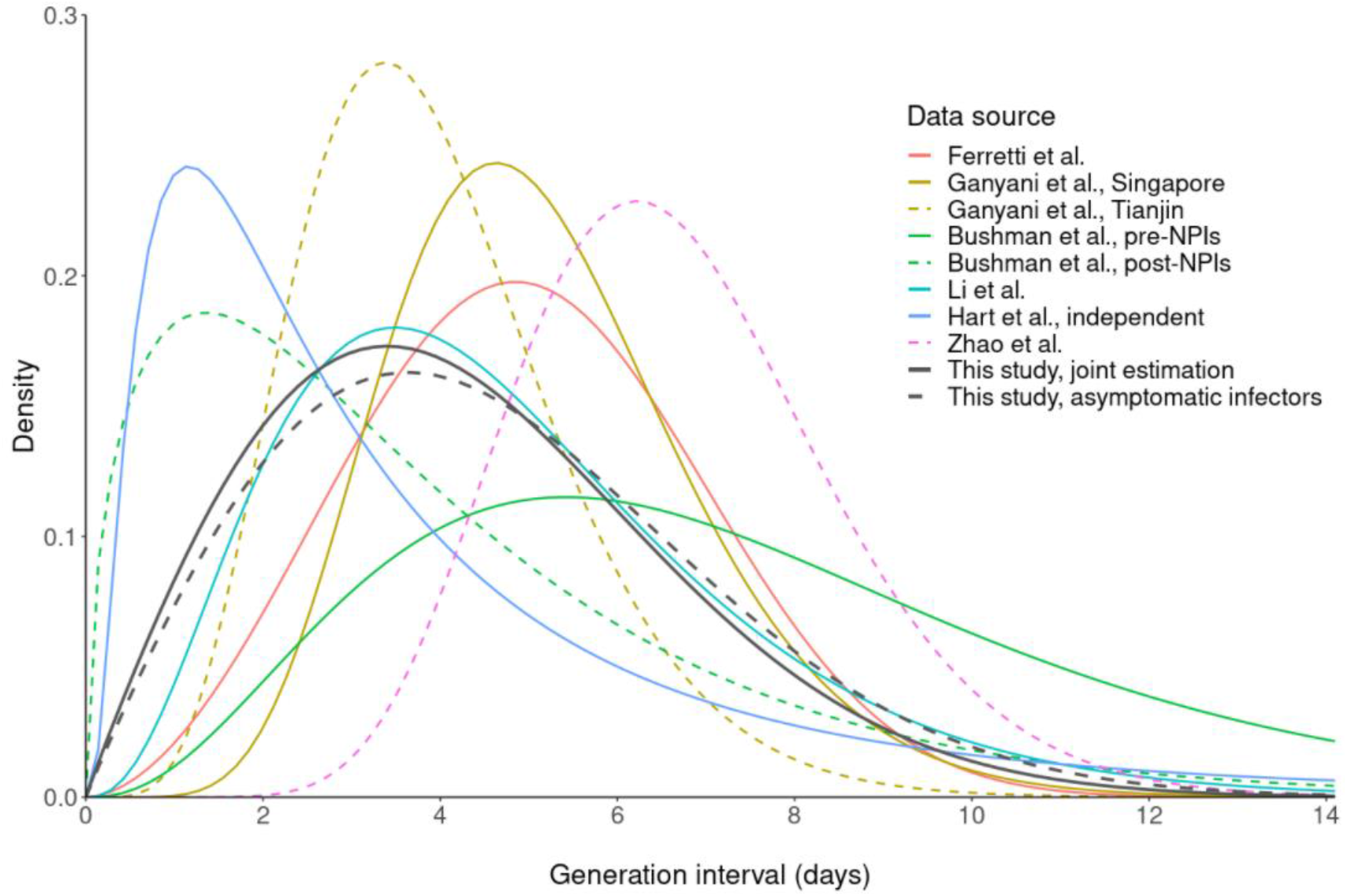
Generation intervals reported across COVID-19 studies. The studies by Ganyani et al., Bushman et al., Li et al., and Zhao et al. are plotted using gamma distributions. The result from Hart et al. was plotted using a lognormal distribution. Results from this study and the study by Ferretti et al. plotted using Weibull distributions.

**Extended Data Fig. 5.**
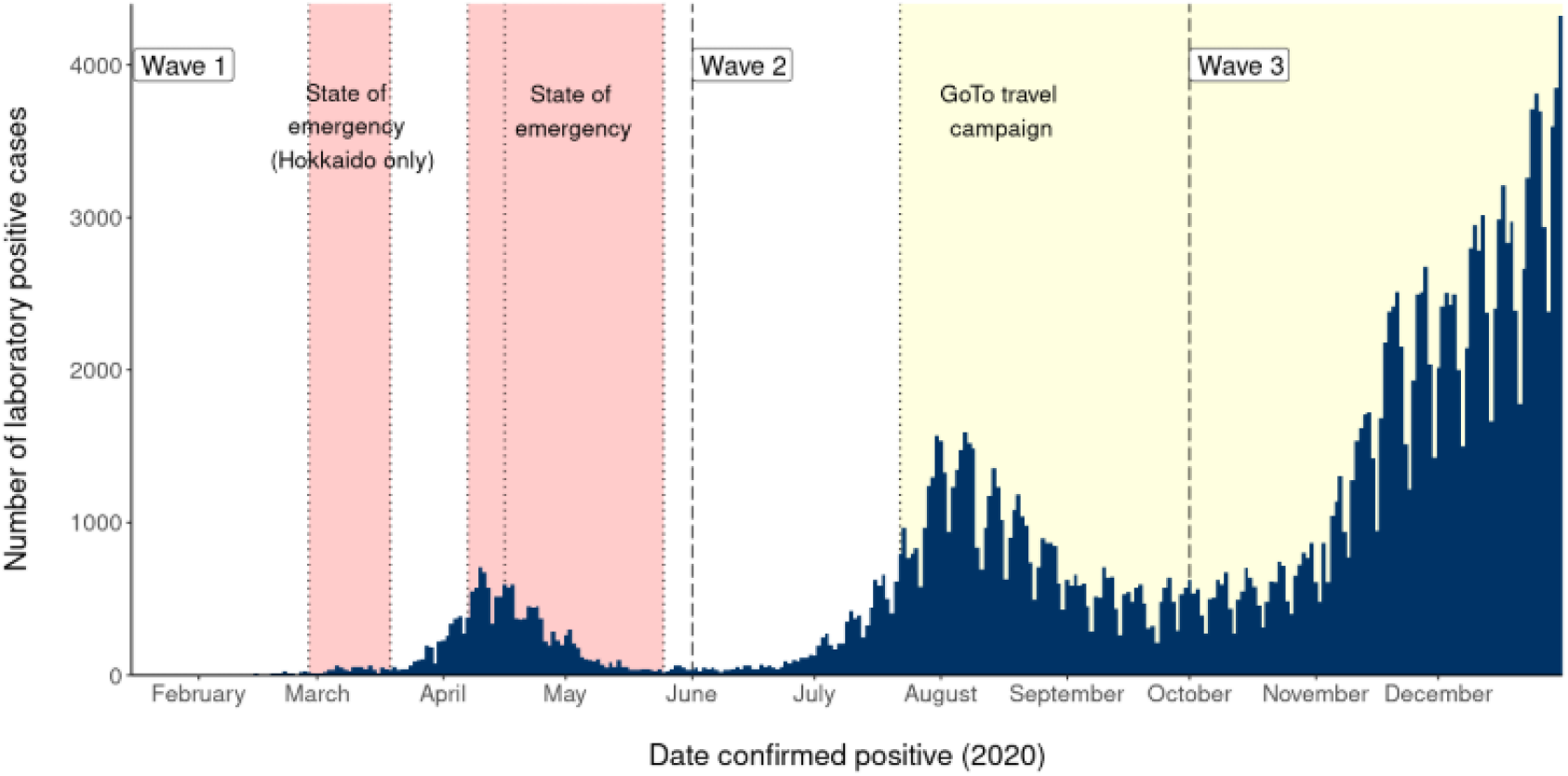
Epidemic curve of COVID-19 cases in Japan by date of laboratory confirmation. Here, the second epidemic wave is shown as beginning 1 June (first dashed line), while the third epidemic wave starts 1 October (second dashed line). Hokkaido declared a local state of emergency 28 February–19 March (first two vertical dotted lines). A national state of emergency was declared for key urban prefectures (Tokyo, Saitama, Chiba, Kanagawa, Osaka, Hyogo, and Fukuoka) on 7 April (third dotted line), with the state of emergency extending nationwide on 16 April (fourth dotted line). The end of the state of emergency varied between prefectures, with most ending 14 May while some continued until 25 May (fifth dotted line). The GoTo travel campaign, offering large discounts on travel inside Japan with the intention of restarting the Japanese economy following the damage caused by COVID-19 related public health and social measures, began on 22 July (sixth dotted line) for all prefectures except Tokyo, which was added to the campaign on 1 October (start date of third wave, second dashed line). The campaign continued through the end of 2020 and into 2021.

