## Supplementary Materials for "Correlation between times to SARS-CoV-2 symptom onset and secondary transmission undermines epidemic control efforts"

**Supplementary information**

1. Previous estimates of the generation interval of COVID-19

Some early estimates of the generation interval of COVID-19 were published during the first half of 2020.[1–3] However, they were not estimated directly from data on exposure dates but derived from estimates of the serial interval and incubation period, as indicated in Supplementary Table 1 below. We found only one other study (Li et al.) that estimated the generation interval directly from reported dates of exposure, although this study assumed infection occurred at the middle of a given exposure period,[4] which may have been intended to address the censored observational data. None of the studies specifically aimed to estimate the generation interval for pairs with asymptomatic infectors.

The studies approached the problem of pair ascertainment (determining directionality of transmission) and estimation of the timing of exposure and contact in different ways.

- Ferretti et al.[1] selected transmission pairs based on high confidence of direct transmission inferred from publicly available sources reported.
- Ganyani et al.[2] used datasets of cases reported in Singapore and Tianjin. For cases linked to clusters, they imputed links between cases to determine transmission pairs and assign directionality.
- Tindale et al.[3] used the same datasets as Ganyani et al. Linkages, when not explicitly available from the data, were established using the methods described in te Beest et al.[5]
- Bushman et al.[6] combined transmission pair data from four published studies.[7–10]
- Li et al.[4] limited their analysis to transmission pairs where the infector had travel history to Hubei Province, China (where the original epicenter of COVID-19, Wuhan, is located). They did not apply doubly-interval censoring, as has been done elsewhere,[11–13] but assumed infection occurred at the exact middle of a given exposure period.
- Hart et al. (worldwide data)[14] combined data from five published studies,[1,7,15–17] and considered a "mechanistic" model where infectors who developed symptoms progressed through different stages of infection.
- Hart et al. (UK data)[18] estimated the generation interval from cases reported in households in the United Kingdom. Their methods considered asymptomatic infectors, and they considered a model where infectiousness and symptom onset were independent, as well as their previously published mechanistic model (see above).
- Zhao et al.[19] used data collected from a previous study that included pairs from China, Japan, and Singapore. The authors used the serial interval and incubation period of the infectee to estimate the generation interval.
- Lau et al.[20] reanalyze data from a previously published study.[21] Their methods do not use the coarse exposure periods to estimate generation interval directly, but assume that symptom onset of the infector is independent of infectiousness conditioning on infection time of the infector.

Among these studies, only Bushman et al. and Hart et al. considered dependence between the generation time and incubation period of the infector. The mechanistic model of Hart et al. considers that symptoms and transmission are not independent, but do not directly quantify possible interdependence. Rather, the model conditions infectiousness on the duration of the incubation period. Bushman et al., although they considered “incubation-dependent” models, found that the best fits for their data were “incubation-independent” models, suggesting low- or no correlation between the generation interval and incubation period. Their method for defining “incubation-dependent” models was to vary the rate parameter of the gamma distribution by dividing it by the length of the incubation period, and it is possible this method—as it did not include correlation as a parameter—could not capture existing correlation. As well, it is possible that the method they used to calculate the generation interval from the serial interval—which assumes independence from the incubation period[1,2,6,22]—does not accurately reflect .

**Supplementary Table 1.** Estimates of the generation interval of COVID-19 from previously published studies

| Estimation method | Mean (95% CI) | SD (95% CI) | Distribution | Geographic scope | Time period | Pairs | Reference |
|---|---|---|---|---|---|---|---|
| SI and IP* | 7.50 (6.81, 8.20) | 3.95 (3.32, 4.74) | Gamma | China, pre-NPI | Jan 2020 | 873† | Bushman[6] |
| SI and IP* | 3.90 (3.59, 4.24) | 3.15 (2.78, 3.53) | Gamma | China, post-NPI | Jan–Feb 2020 | 873† | Bushman |
| SI and IP | 4.83 (4.31, 5.40) | 1.73 (0.98, 2.55) | Gamma | Worldwide | Jan–Mar 2020 | - | Challen[23] |
| SI and IP* | 5.04 (4.19, 6.31) | 1.93 (1.52, 2.47) | Weibull | Worldwide | Dec 2019–Feb 2020 | 40 | Ferretti[1] |
| SI and IP‡ | 5.20 (3.78, 6.78) | 1.72 (0.91, 3.93) | Gamma | Singapore | Jan–Feb 2020 | 54 | Ganyani[2] |
| SI and IP‡ | 3.95 (3.01, 4.91) | 1.51 (0.74, 2.97) | Gamma | Tianjin, China | Jan–Feb 2020 | 45 | Ganyani |
| Mechanistic* | 5.57 (5.08, 6.09) | 2.32 (1.83, 2.91) | Gamma | Worldwide | Dec 2019–Mar 2020 | 191 | Hart[14] |
| Independent§ | 4.2 (3.3, 5.3) | 4.9 (3.0, 8.3) | Lognormal | United Kingdom | Mar–Nov 2020 | ¶ | Hart[18] |
| Mechanistic§ | 6.0 (5.2, 7.0) | 4.9 (4.0, 6.3) | Gamma | United Kingdom | Mar–Nov 2020 | ¶ | Hart |
| Exposure and onset times | 5.7 (4.8, 6.5) | 1.7 (0.7, 2.5) | Gamma | China | Jan 2020 | 81** | Lau[20] |
| Exposure times | 4.81 (4.13, 5.58) | 2.52 (1.93, 3.32) | Gamma | China | Jan–Feb 2020 | 67 | Li[4] |
| IP intermediates | 3.71 (2.36, 4.91) | - | Gamma | Singapore | Jan–Feb 2020 | 56** | Tindale[3] |
| IP intermediates | 2.82 (1.82, 3.52) | - | Gamma | Tianjin, China | Jan–Feb 2020 | 72** | Tindale |
| SI and infectee IP | 6.7 (5.4, 7.6) | 1.8 (0.3, 3.8) | Gamma | China, Japan, Singapore | Dec 2019–Apr 2020 | 254 | Zhao[19] |

CI: confidence/credible interval; COVID-19: coronavirus disease 2019; IP: incubation period; NPI: non-pharmaceutical intervention; SD: standard deviation; SI: serial interval. *IP distribution based on Lauer et al.[24] †Total pairs for both pre- and post-NPI. ‡IP distribution based on Zhang et al.[10] §IP distribution based on McAloon et al.[25] ¶172 households with 603 cases. **Manually counted from figure.

Supplementary Table 2 lists previously published studies where correlation between transmission intervals (generation or serial interval) and the incubation period were assessed. Dependence between the two parameters has rarely been estimated.

**Supplementary Table 2.** Estimates of correlation between transmission intervals and incubation periods from other studies

| Disease | Transmission interval | Kendall's tau (CrI or p-value) | Geographic scope | Timeframe | Method | Reference |
|---|---|---|---|---|---|---|
| Measles | GI | 0.27 (-0.34, 0.76)* | Rhode Island | 1917–1923 | Copula, gamma | Klinkenberg[26] |
| Measles | GI | 0.63 (0.14, 0.88)* | Rhode Island | 1929–1934 | Copula, gamma | Klinkenberg |
| Measles | GI | 0.72 (0.52, 0.84)* | Rhode Island | 1917–1923 | Copula, lognormal | Klinkenberg |
| Measles | GI | 0.84 (0.67, 0.95)* | Rhode Island | 1929–1934 | Copula, lognormal | Klinkenberg |
| COVID-19 | SI† | 0.13 (p=0.2) | Singapore | 2020 | Case pairs | Tindale[3] |
| COVID-19 | SI† | 0.19* (-) | Tianjin, China | 2020 | Case pairs | Tindale |

CrI: credible interval; COVID-19: coronavirus disease 2019; GI: generation interval; SI: serial interval. *Original results were presented as correlation coefficient $\rho$, which is defined in terms of Kendall's tau as $\tau_K = \frac{2}{\pi}\arcsin(\rho)$. †Estimates of the serial interval included only positive values.

Klinkenberg and Nishiura[26] estimated the correlation between the generation interval and incubation period of measles in Rhode Island prior to development of the measles vaccine. They found very different results depending on whether they were using bigamma or bilognormal marginals. The better fit to the bilognormal marginals may reflect the better fit to the lognormal distribution commonly seen in incubation period data for respiratory diseases,[27] including COVID-19.[12,13,24] Tindale et al. assessed covariance between the serial interval and incubation period of COVID-19 using empirical data from their selected transmission pairs. However, the pairs ignored negative serial intervals.

### 2. Case definition and determination of transmission pairs

Nearly all COVID-19 cases in Japan had a positive viral test for SARS-CoV-2, with a few exceptions made for cases with positive antibody tests based on clinical judgement. These viral tests were either nucleic acid amplification tests (NAATs) or antigen tests. Not all public health jurisdictions shared the type of test in their case reports, but typically the NAATs were either reverse transcription polymerase chain reaction (RT-PCR) or loop-mediated isothermal amplification (LAMP) tests.

Presymptomatic and asymptomatic transmission are possible for SARS-CoV-2 infections.[3,7] Thus, directionality of transmission for epidemiologically linked cases that is determined solely based on dates of onset among linked cases is likely to include some misclassification of infector and infectee status, and may ignore possible intermediate infectors. We collected information on COVID-19 cases reported in Japan and looked for epidemiological information (i.e. contact and travel history) that would provide insight into directionality of transmission between linked cases. Criteria for ascertainment of directionality of transmission generally fell into one of four categories:

1. Import: If a potential infector had onset during or following international travel to a country with COVID-19 cases they were considered an imported case.
2. Cluster: A cluster was defined as five or more cases. If a potential infector was linked directly or via a chain of infections to a cluster (common exposure) the link was classified as cluster-based.
3. Domestic travel: If onset of an infector occurred during or within 10 days after travel to another prefecture and there were no more obvious possible exposures these cases were labelled as having domestic travel as their possible exposure, and the dates of travel form the left- and right-hand bounds of their exposure.
4. Contact pattern: the contact pattern between cases, typically supplemented by some reported dates of contact or exposure, provided insight into directionality of infection for linked infector-infectee pairs.

We supply a variable "link_basis" in the dataset of transmission pairs as a means of assessing whether there was any difference between estimates given the differing rationales for ascertaining directionality of transmission for pairs for each of the above categories.

3. Cleaning dates of exposure and contact

Reported dates on exposure, contact, and symptom onset were cleaned to obtain EL and ER (left- and right-hand exposure times for the infector), CL and CR (left- and right-hand times of contact between the infector and infectee), as well as S1 and S2 (symptom onset times of the infector and infectee). All times are reported as number of days relative to a "time zero" of 1 January 2020. An abridged data dictionary for calculated values is provided in

Supplementary Table 3.

For most case reports that included temporal information on potential exposures or contact between cases, exact dates or date ranges were explicitly stated. However, some of the data may include inferences made from statements related to parts of the month, with data entry performed as follows:

- Beginning of the month = until the seventh day of the month. More common may be just the first day of the month, or until the third day.
- End of the month = from the 25th day of the month. An informal survey found that approximately one-third of respondents felt that the phrase “end of the month” could include dates as early as the 25th (see https://mainichi-kotoba.jp/enq-255).
- Beginning, middle, and end of the month = days 1–10, 11–20, 21–last day of a given month, respectively.

In cases where left- or right-hand bounds for exposure and contact were missing from the data, we used other epidemiological information to substitute for these bounds when plausible to do so. The assumptions were as follows:

1) Data cleaning applied to ER/CR:
    a) For some cases, recorded right-hand exposure (e.g., travel, contact with a case) for infector and/or infectee may exceed symptom onset (S1 or S2). As we are only interested in exposure and contact dates as they related to the possible infection time for infector and infectee, we set ER and CR equal to S1 and S2.
    b) Similarly, for some infectors, their recorded right-hand exposure may exceed their time of contact with the infectee. In these scenarios we presume the infector is infected by the time of final contact the infectee, so we censor ER to CR.
    c) Missing ER were sequentially assigned to the minimum of either:
        i) S1,
        ii) CR,
        iii) date of infector laboratory confirmation,
        iv) date of infector isolation,
        v) or 14 days* after EL.
    d) Missing CR were sequentially assigned to the minimum of either:
        i) S2,
        ii) date of infectee laboratory confirmation,
        iii) or 14 days* after CL.
2) Data cleaning applied to EL/CL:
    a) Missing EL were assigned to 14 days* before ER.
    b) In some cases, infectees may have been reported to have contact with the infector prior to the infector’s left-hand exposure (EL). For example, the infector and infectee may have met several

times. It is possible that the left-hand exposure for the infectee (CL) was assigned to a date that was earlier than EL. However, as pairs included in the dataset were selected for having a relatively high likelihood of the directionality of infection being true, in cases where CL < EL, we set CL = EL.

c) If the infector exposure type was labelled “International travel” and the infectee was missing CL, then CL was set to ER.

d) If contact type between infector and infectee is “household,” link basis was “cluster” or “domestic contact”, infectee was missing CL, and EL was not missing, CL was set to EL.

e) Missing CL were assigned to EL, as an infector cannot be infectious (and therefore CL cannot be a valid date of contact for transmission to occur from infector to infectee) unless the infector’s own transmission has occurred.

*14 days was selected as this ~>95% of the incubation period.[12,13,24] Although the above assumptions made it possible to obtain EL and ER even in a left- or right-hand bound of exposure was not reported, we only included cases where a left- or right-hand bound of exposure was explicitly reported.

**Supplementary Table 3.** Calculated variables included in the dataset

| Variable | Description | Class | Values | Details |
|---|---|---|---|---|
| EL | Left-hand bound of infector exposure | Numeric | Days | |
| ER | Right-hand bound of infector exposure | Numeric | Days | |
| CL | Left-hand bound of infectee contact with infector | Numeric | Days | |
| CR | Right-hand bound of infectee contact with infector | Numeric | Days | |
| S1 | Symptom onset day of infector in days | Numeric | Days | |
| S2 | Symptom onset day of infectee in days | Numeric | Days | |
| link_basis | What the basis was for determining directionality of transmission between linked pairs | Factor | Cluster | Infector was linked to a cluster or part of a transmission chain linked back to a cluster |
| | | | Contact pattern | Timings of contact and onset between cases provide plausible evidence for directionality of transmission |
| | | | Domestic travel | Infector travelled domestically to a location with ongoing transmission and timing of travel is plausibly related to onset of disease |
| | | | International travel | Infector travelled internationally part of a transmission chain linked back to international travel, and said travel is believed to have been the source of infection |
| exposure_type | Transmission setting for infector exposure. | Factor | Household | A household member or family member (when household status was not specified) |
| | | | Social-contact based interaction | Venue for interaction is based on social interaction. Restaurants, nightlife, karaoke, sports events, live music, gyms, friends, relatives, acquaintances, etc., or type of contact is not specified. |
| contact_type | Transmission setting for contact between infector and infectee. | | Core community interaction | Venue for exposure are schools, general workplaces, essential workplaces (care facilities, medical facilities, government services, etc.), or exposure is related to travel to another area and source of infection is unknown (community infection). |

4. Bivariate joint distribution

We employed a Bayesian approach combining copulas (multivariate cumulative distribution functions) with doubly-interval censoring to obtain estimates of the generation interval and incubation period, as well as a measure of correlation those two parameters. For $i \in \{1, \dots, N\}$ transmission pairs, we obtain the following doubly-interval censored likelihoods for the generation interval and incubation period for data $D$:

$$L1(\Theta_j; \mathrm{D}) = \prod_i \int_{E_{L.i}}^{E_{R.i}} \int_{C_{L.i}}^{C_{R.i}} j(e) f(\mathrm{c} - e) \mathrm{dcd}e, \quad (1)$$

$$L2(\Theta_j; \mathrm{D}) = \prod_i \int_{E_{L.i}}^{E_{R.i}} \int_{S_{L.i}}^{S_{R.i}} j(e) g(\mathrm{s} - e) \mathrm{dsd}e.$$

Here, $e$ is the time of infection of the infector and $j(.)$ is the probability distribution function (PDF) of the time of infection of the infector following a uniform distribution across exposure time $E_L$ to $E_R$. Similarly, $c$ is the time of transmission of the pathogen from infector and infectee occurring between $C_L$ and $C_R$, with $f(.)$ representing the PDF of the generation interval. Finally, $s$ is the time of symptom onset of the infector occurring between $S_L$ and $S_R$, with $g(.)$ representing the PDF of the incubation period.

Combining doubly-interval censoring with a copula function allowed us to obtain the bivariate joint distribution of the generation interval and incubation period. Copulas provide a correlation structure to sets of marginal distributions, allowing for the marginal distributions and dependence structure to be modeled separately. In accordance with Sklar's theorem,[28] the joint cumulative distribution function (CDF) can be decomposed into the copula and univariate marginal CDFs. As such, if $H(x, y)$ is a joint bivariate CDF with marginal CDFs $F(x)$ and $G(y)$, there exists a copula $C$: $[0,1]^2 \longrightarrow [0,1]$ such that:

$$H(x, y) = C\big(F(x), G(y)\big) \quad (2)$$

for all $(x, y) \in [-\infty, \infty]^2$. From relations $F(x) = u$ and $G(y) = v$ we obtain $x = F^{-1}(u)$ and $y = G^{-1}(v)$. Substituting $F^{-1}(u)$ and $G^{-1}(v)$ into (2) we obtain the copula:

$$C(u, v) = H(F^{-1}(u), G^{-1}(v)), \quad (3)$$

for all $(u, v) \in [0,1]^2$. The marginal distribution functions are given by $F(x) = \int_{-\infty}^{x} f(u) du$ and $G(y) = \int_{-\infty}^{y} g(v) dv$. From the marginal distributions we have $U = F(X)$ and $V = G(Y)$ which are uniformly distributed in [0,1]. The joint distribution of $(U, V)$ is the copula $C(u, v) = P(U \leq u, V \leq v)$. The density of the bivariate copula $C$ is:

$$c(u, v) = \frac{\partial^2 C(u, v)}{\partial u \partial v} \quad (4)$$

Setting $x$ and $F(x)$ to be the data and CDF of the generation interval, and $y$ and $G(y)$ to be the data and CDF of the generation interval, the joint distribution described in equation (2) is therefore the joint distribution of the incubation period and generation interval. Given a copula parameter $\theta$, the overall log-likelihood is expressed as the sum of the log-likelihood of each marginal distribution plus the log-likelihood of the copula:

$$\log L(\mu_X, \sigma_X, \mu_Y, \sigma_Y, \theta | x, y) = \log L(\mu_X, \sigma_X | x) + \log L(\mu_Y, \sigma_Y | y) + \log L(\theta | u, v). \quad (5)$$

*Copula selection*

The Gaussian copula was utilized by Klinkenberg and Nishiura to assess correlation between the generation interval and incubation period for measles data,[26] and therefore was of interest for inclusion in this study. However, the Gaussian copula does not account for tail dependence. To consider lower tail dependence we included the Clayton copula, while to consider upper tail dependence we included the Gumbel copula. The independence copula, which is the copula that results from a dependency structure in which each individual variable is independent of each other, was also considered. Supplementary Table 4 introduces various properties of these four copulas.

*Gaussian copula*

The Gaussian copula is an elliptical copula. It is defined by

$$C(u, v) = \Phi_\theta\left(\Phi^{-1}(u), \Phi^{-1}(v)\right), \quad (6)$$

where $\Phi$ is the standard normal distribution function and $\theta \in (-1,1)$ is the correlation between the components. The density of the bivariate Gaussian copula is given by

$$c(u, v; \theta) = \frac{1}{\sqrt{1-\theta^2}} exp\left\{-\frac{\theta^2(s^2+t^2) - 2\theta st}{2(1-\theta^2)}\right\}, \quad (7)$$

where $s = \Phi^{-1}(u)$ and $t = \Phi^{-1}(v)$. As such, the log-likelihood function for the Gaussian copula is

$$\log\left(L(\rho | u, v)\right) = -\frac{1}{2}\log(1-\theta^2) - \frac{\theta^2(s^2+t^2) - 2\theta st}{2(1-\theta^2)}. \quad (8)$$

For Gaussian copula, Kendall's tau is defined as $\frac{2}{\pi}\sin^{-1}(\theta)$. Independence is reached when $\theta = 0$.

*Independence, Gumbel, and Clayton copulas*

The Gumbel and Clayton copulas are single-parameter Archimedian copulas.[29] As a family, Archimedian copula are defined as

$$C(u, v) = \varphi^{[-1]}\left(\varphi(u) + \varphi(v)\right), \quad (9)$$

where $\varphi$ is the generator function of the copula $C$. $\varphi: [0,1] \rightarrow [0, \infty]$ is a continuous strictly decreasing convex function such that $\varphi(1) = 0$ and $\varphi^{[-1]}$ is the pseudo-inverse of $\varphi$, defined as $\varphi^{[-1]}: [0, \infty] \rightarrow [0,1]$ with

$$\varphi^{[-1]}(t) = \begin{cases} \varphi^{-1}(t), & 0 \le t \le \varphi(0), \\ 0, & \varphi(0) \le t \le \infty. \end{cases} \tag{10}$$

For the bivariate Gumbel copula, the generator is $\varphi(t) = (-log(t))^{\theta}$ and inverse generator $\varphi(t)^{-\theta} = \exp(-t^{1/\theta})$ for copula parameter $\theta \in [1, \infty)$, where $t$ varies from 0 to 1 regardless of whether it is equal to $u$ or $v$. The bivariate Gumbel copula is given as

$$C(u,v) = exp\left\{-\left[(-\ln(u))^{\theta} + (-\ln(v))^{\theta}\right]^{\frac{1}{\theta}}\right\}, \qquad \theta \in [1, \infty), \tag{11}$$

where $\theta \to \infty$ indicates full dependence, while $\theta = 1$ corresponds to independence. The Gumbel copula has upper-tail dependence and is useful for datasets where the dependence between high values of the univariate distributions is stronger than the dependence between their low values. The Gumbel copula does not allow negative dependence. Its density is given by

$$c(u,v;\theta) = \frac{C(u,v)}{(uv)}(\log(u)\log(v))^{\theta-1} w^{\frac{2}{\theta}-2}\left(1 + (\theta-1)w^{\frac{1}{\theta}-2}\right), \tag{12}$$

where $w = (-log(u))^{\theta} + (-log(v))^{\theta}$. Consequently, the log-likelihood function[30] is defined by

$$\log\big(L(\theta|u,v)\big) = w^{\frac{1}{\theta}} - \log(uv) + (\theta-1)\log\big(\log(\mathrm{u})\log(\mathrm{v})\big) \\ + \log\left(w^{\frac{2}{\theta}-2}\right)\log\left[(\theta-1)w^{\frac{1}{\theta}-2}\right]. \tag{13}$$

For the bivariate Clayton copula, the generator is $\varphi(t) = \frac{1}{\theta}\left(t^{-\theta} - 1\right)$ and inverse generator $\varphi(s)^{-\theta} = \max\left\{(1+\theta s)^{-1/\theta}, 0\right\}$ for $\theta \in [-1, \infty)\backslash\{0\}$. The bivariate Clayton copula is given as

$$C(u,v) = max\left(\left(u^{-\theta} + v^{-\theta} - 1\right)^{-\frac{1}{\theta}}, 0\right), \tag{14}$$

where full dependence is reached as $\theta \to \infty$, while independence is reached as $\theta \to 0$.

The density is given by

$$c(u,v;\theta) = \frac{(\theta+1)(uv)^{\theta}}{(u^{\theta}v^{\theta}(uv)^{\theta})^{\frac{1}{\theta}+2}} \tag{15}$$

The log-likelihood function for the Clayton copula is defined by

$$\log\big(L(\theta|u,v)\big) = \log(1) + \theta - (\theta+1)(\log(u) + \log(v)) \\ - \left(\frac{1+2\theta}{\theta}\right)\log\left(u^{-\theta} + v^{-\theta} + 1\right) \tag{16}$$

The independence copula is a special case of several Archimedean copulas, as well as a special case of the Gaussian copula with a correlation matrix equal to the identity matrix. It has no correlation (copula) parameter and no tail dependence. For the bivariate independence copula, the generator is $\varphi(t) = \exp(-t)$. We applied the independence copula as the special case of the Gumbel copula ($\theta = 1$).

*Measuring dependence*

We use Kendall's tau to assess dependence between the generation interval and incubation period. Rank correlations such as Kendall's tau only depend on the unique copula of the joint distribution and are therefore invariant to monotone transformations of the marginals.[29] The relationship between Kendall's tau and the various copula parameters are listed in Supplementary Table 4.

**Supplementary Table 4.** Bivariate copulas and their properties

| Copula | Copula parameter | Independence | Kendall's tau | Range of tau | Lower tail dependence | Upper tail dependence |
|---|---|---|---|---|---|---|
| Gaussian | $\theta \in (-1,1)$ | $\theta = 0$ | $\frac{2}{\pi}\sin^{-1}(\theta)$ | [-1,1] | 0 | 0 |
| Gumbel | $\theta \in [1,\infty)$ | $\theta = 1$ | $1 - \theta^{-1}$ | [0,1] | 0 | $2 - 2^{\frac{1}{\theta}}$ |
| Clayton | $\theta \in [-1,\infty)\backslash\{0\}$ | $\theta \to 0$ | $\frac{\theta}{\theta + 2}$ | [0,1] | $2^{-\frac{1}{\theta}}$ | 0 |
| Independence | None | Always | 0 | 0 | 0 | 0 |

Of the three copula that allow for dependence between the parameters (Gaussian, Gumbel, Clayton), only the Gaussian copula considers negative correlation.

*Mixture model*

We used a Bayesian mixture model to determine the best-fit combination of various copulas and marginal distributions. For all possible combinations of copula and marginal distributions we assign individual likelihoods as the sum of component contributions and formulated the model in terms of latent variables. We considered the four copulas described above, and the gamma, lognormal, and Weibull distributions for the generation interval and incubation period for a total of $M = 4 \times 3 \times 3 = 36$ combinations. These $M$ combinations mix in proportion $\lambda$, where $\lambda_m \geq 0$ and $\sum_m^M \lambda_m = 1$.

The outcome is drawn from one of these combinations, the identity of which is controlled by a categorical mixing distribution $z \sim \text{Categorical}(\lambda)$. We fixed Kendall's tau and the means and SDs for the generation interval and incubation period across all possible combinations $M$. We used informative priors for the means of the generation interval and incubation period, obtained from previous publications.[13,31]

**Author contributions**

NML and NH conceived the study. Data were collected by NML. Data were analyzed by NML and ARA, and interpreted by NML, ARA, and HN. NML wrote the first draft of the manuscript, which was revised by NML, ARA, and HN. All authors approved the final version of the manuscript.

**Competing interests**

The authors declare no competing interests.

**Acknowledgements**

The authors thank the reporting jurisdictions in Japan for conducting thorough investigations and publishing information about cases and transmission settings of public health import, Atsuna Tokumoto for help with initial data collection on transmission links for two prefectures, and the Nishiura lab for additional support with data collection on case characteristics. NML received a Japanese Ministry of Education, Culture, Sports, Science, and Technology (MEXT) graduate scholarship. HN received funding from a Health and Labor Sciences Research Grant (20CA2024 and 20HA2007), the Japan Agency for Medical Research and Development (AMED; JP20fk0108140 and JP20fk0108535s0101), the Japan Society for the Promotion of Science (JSPS)KAKENHI (21H03198), and the Japan Science and Technology Agency (JST) SICORP (e-ASIA) program (JPMJSC20U3).
